# The anti-SARS-CoV-2 immunoglobulin G levels and neutralising capacities against alpha and delta virus variants of concern achieved after initial immunisation with vector vaccine followed by mRNA vaccine boost are comparable to those after double immunisation with mRNA vaccines

**DOI:** 10.1101/2021.07.09.21260251

**Authors:** Ruben Rose, Franziska Neumann, Olaf Grobe, Thomas Lorentz, Helmut Fickenscher, Andi Krumbholz

## Abstract

**Background:** The humoral immune response after primary immunisation with a SARS-CoV-2 vector vaccine (AstraZeneca AZD1222, ChAdOx1 nCoV-19, Vaxzevria) followed by an mRNA vaccine boost (Pfizer/BioNTech, BNT162b2; Moderna, m-1273) was examined and compared with the antibody response after homologous vaccination schemes (AZD1222/AZD1222 or BNT162b2/BNT162b2).

**Methods:** Sera from 59 vaccinees were tested for anti-SARS-CoV-2 immunoglobulin G (IgG) and virus-neutralising antibodies (VNA) with four IgG assays, a surrogate neutralisation test (sVNT), and a Vero cell-based neutralisation test (cVNT) using the B.1.1.7 variant of concern (VOC; alpha) as antigen. Investigation was done before and after heterologous (n=31 and 42) or homologous booster vaccination (AZD1222/AZD1222, n=8/9; BNT162b2/BNT162b2, n=8/8). After the second immunisation, 26 age and gender matched sera (AZD1222/mRNA, n=9; AZD1222/AZD1222, n=9; BNT162b2/BNT162b2, n=8) were also tested for VNA against VOC B.1.617.2 (delta) in the cVNT. The strength of IgG binding to separate SARS-CoV-2 antigens was measured by avidity.

**Results:** After the first vaccination, prevalence of IgG directed against (trimeric) SARS-CoV-2 spike (S)-protein and its receptor-binding domain (RBD) varied from 55-95 % (AZD1222) to 100% (BNT162b2), depending on the vaccine used and the SARS-CoV-2 antigen used. The booster vaccination resulted in 100 percent seroconversion and appearance of highly avid IgG as well as VNA against VOC B.1.1.7. The results of the anti-SARS-CoV-2 IgG tests showed an excellent correlation to the VNA titres against this VOC. The agreement of cVNT and sVNT results was good. However, the sVNT seems to overestimate non and weak B.1.1.7-neutralising titres. The mean anti-SARS-CoV-2 IgG and B.1.1.7-neutralising titres were significantly higher after heterologous vaccination compared to the homologous AZD1222 scheme. If VOC B.1.617.2 was used as antigen, significantly lower mean VNAs were measured in the cVNT, and three (33.3%) vector vaccine recipients had a VNA titre <1:10.

**Conclusions:** The heterologous SARS-CoV-2 vaccination leads to a strong antibody response with anti-SARS-CoV-2 IgG and VNA titres at a level comparable to that of a homologous BNT162b2 vaccination scheme. Irrespectively of the chosen immunisation regime, highly avid IgG antibodies can be detected just two weeks after the second vaccine dose indicating the development of a robust humoral immunity. The observed reduction in the VNA titre against VOC B.1.617.2 is remarkable and may be attributed to a partial immune escape of the delta variant.

## Background

Since spring 2020 the pandemic caused by the *severe acute respiratory syndrome coronavirus 2* (SARS-CoV-2) [1] is ongoing and represents a global challenge. The availability of safe and effective vaccinations are seen as one of the most important pillars in containing the pandemic [2, 3]. Within a few months, the intensive research activities led to the development of several highly effective SARS-CoV-2 vaccines [4, 5, 3]. In addition to the induction of cellular immunity, their administration should stimulate the formation of virus-neutralising antibodies (VNA) that bind to epitopes of the viral spike (S)-protein and its receptor binding domain (RBD) and, thus, prevent the cells from being infected [6, 7, 3].

Four SARS-CoV-2 vaccines have received conditional approval in the European Union. These vaccines are based on two different technologies [8]. For the messenger ribonucleic acid (mRNA) vaccines from Pfizer/BioNTech (BNT162b2) and Moderna (mRNA-1273), the genetic information for the S-protein was genetically optimised and the mRNA was packaged in liposomes. After inoculation, the muscle cells directly expressed this stable and highly immunogenic viral surface protein [2, 6]. In vector vaccines, replication-deficient human (Ad26.COV2; Janssen) or chimpanzee adenoviruses (ChAdOx1 nCoV-19/AZD1222, Vaxzevria; AstraZeneca, hereinafter referred to as AZD1222) are used to introduce the genetic information of the SARS-CoV-2 S-protein into the cells, followed by transcription of deoxyribonucleic acid into mRNA and expression of the S-protein [2, 6].

Due to the widespread use of these vaccines, rare and sometimes unexpected side effects have been reported. Particularly noteworthy are cases of immune thrombotic thrombocytopenia, which predominantly occurred in women under 50 years of age within one month after the initial vaccination with AZD1222 [5]. Many of these patients developed cerebral sinus venous thrombosis or splanchnic vein thrombosis and presented antibodies to platelet factor 4 but without previous exposure to heparin [5]. Due to this rare but serious side effect, AZD1222 is no longer unreservedly recommended by the Standing Vaccination Commission (STIKO) of the Robert Koch Institute for individuals under 60 years of age. The STIKO suggests that a vaccination with AZD1222 that has already started should be completed with an mRNA vaccine [9, 10]. Due to the sharp increase in the delta variant of concern (VOC; Pango-lineage [11] B.1.617.2) in Germany, the STIKO has revised its recommendations once more. Since July 1^st^ 2021, all AZD1222 first vaccinated persons have been recommended to complete the second vaccination with an mRNA vaccine [12]. Animal experiments indicated very good humoral and cellular immunity after heterologous vaccination [13, 14]. There is, however, so far limited knowledge on the benefit of the heterologous vaccination scheme in humans. First results indicated a higher prevalence of short-lived side effects following the heterologous boost dose compared to the homologous counterpart [15]. Meanwhile, a few studies have been published for the immunogenicity of the AZD1222/mRNA vaccine regimen [16-21].

In this report, we compare the SARS-CoV-2 specific immunoglobulin G (IgG) response after heterologous immunisation with that elicited by homologous vaccination schedules. We also focus on the developing anti-SARS-CoV-2 IgG avidity as a parameter for IgG maturity and binding strength. Finally, we use various methods to investigate the development of VNA against two prevalent VOCs. Therefore, humoral immunity induced by vaccines is being extensively studied. We believe that the results obtained in this study will help to better understand the effects and possibly benefits of a heterologous vaccination regimen.

## Methods

The anti-S and anti-RBD IgG response after heterologous immunisation with a SARS-CoV-2 vector vaccine as prime and an mRNA vaccine as boost was compared to that after homologous vaccination with vector or mRNA vaccines. This setting also includes monitoring of IgG avidity and of virus-neutralising capacities. Forty-seven female and twelve male vaccinees with a median age of 31 years (age span 18 – 61 years) were recruited for this study and gave their informed consent. Forty-two of them received a heterologous immunisation scheme (N=41, AZD1222/BNT162b2; N=1, AZD1222/mRNA-1273), while nine and eight vaccinees received a homologous scheme of the vector vaccine AZD1222 or the mRNA vaccine BNT162b2, respectively. The first blood sample was taken immediately before the second vaccination and the second about two weeks later. For several individuals no serum was available before or after the booster vaccination (Table 1). The ethics committee of the medical faculty of the Christian-Albrechts-Universität zu Kiel (Kiel, Germany) approved the study design (D467/20, 16.04.2020, amendment 02.02.2021). We examined the early humoral immune response (other samples obtained a few days to weeks after the initial immunisation with AZD1222) of most of the subjects in a previous study. In addition, sera obtained from three vaccinees after the initial immunisation with BNT162b2 (N=2) and AZD1222 (N=1) have already been tested in frame of the previous study [22]. In this respect, we consider it justified to include these individuals (and few sera) in the present report and to demonstrate the results before and after the booster vaccination.

**Table 1.** Individuals included in this study

| Study groups | Number of individuals after 1<sup>st</sup>/2<sup>nd</sup> vaccination | Median age in years | Age span in years | Gender (female/male) | Time (median) from 1<sup>st</sup>/2<sup>nd</sup> vaccination up to 1<sup>st</sup>/2<sup>nd</sup> serum sampling in days |
| --- | --- | --- | --- | --- | --- |
| *Heterologous vaccination scheme* |  |  |  |  |  |
| AZD1222/BNT162b2 | 30/41 | 27 | 18-56 | 33/8 | 69/16 |
| AZD1222/BNT162b2<sup>†</sup> | -/9 | 43 | 23-56 | 7/2 | -/14 |
| AZD1222/mRNA-1273 | 1/1 | 45 | 45-45 | 1/0 | 64/14 |
| *Homologous vaccination scheme* |  |  |  |  |  |
| AZD1222<sup>†</sup> | 8/9 | 41 | 23-61 | 6/3 | 69/16 |
| BNT162b2<sup>†</sup> | 8/8 | 35 | 23-51 | 7/1 | 34/14 |

<sup>†</sup> After the second vaccination, several sera were also tested for presence of virus-neutralising antibodies against the SARS-CoV-2 delta variant of concern (B.1.617.2). For this purpose, individuals from the group of heterologous vaccinations whose age and gender largely corresponded to those with homologous vaccinations were chosen.

### Anti-SARS-CoV-2 specific IgG immunoassays

The sera were tested with the SERION ELISA agile SARS-COV-2 IgG assay (S-protein as antigen; Institut Virion\Serion GmbH, Würzburg, Germany) and the Abbott SARS-CoV-2 IgG II Quant assay (RBD as antigen; Abbott, Wiesbaden, Germany) as described previously [22]. In addition, the LIAISON® SARS-CoV-2 Trimeric S IgG assay was included as a further assay on a LIAISON® XL system (both Diasorin S.p.A, Saluggia, Italy). According to the manufacturer, this quantitative chemiluminescence immunoassay detects IgG directed against the trimeric S-protein and has an excellent clinical sensitivity and specificity of 98.7% and 99.5%, respectively. The high diagnostic value of this test has also been demonstrated in a recent seroprevalence study [23]. The results of the three IgG assays were given in Binding Antibody Units (BAU) per milliliter (ml), using the manufacturer’s conversion factors, which were based on measurements of the WHO International Standard Anti-SARS-CoV-2 Immunoglobulin (NIBSC code 20-136) [24]. As in our previous studies, we generally rate borderline test results as positive [25, 22].

### Anti-SARS-CoV-2 IgG immunoblots including measurement of IgG avidities

All sera were tested with and without avidity reagent in the *recom*Line SARS-CoV-2 IgG assay (Mikrogen GmbH, Neuried, Germany) as reported previously [25, 22]. The strength of SARS-CoV-2 antigen binding against the S1-and RBD-subunits of the S-protein and against the nucleoprotein (NP, if detectable) was assigned to the categories low (= 1), intermediate (= 2), and high (= 3) [22].

### Measurement of SARS-CoV-2-neutralising antibodies

The sera were examined for their virus-neutralising capacities. First, a surrogate assay was used (TECO® SARS-CoV-2 Neutralisation Antibody ELISA; TECOmedical AG, Sissach, Switzerland). In this competetive assay, the human angiotensin-converting enzyme 2 (ACE-2) was attached to the solid phase while peroxidase-conjugated RBD was present in the liquid phase. If the human serum contained RBD-specific antibodies, binding of RBD to ACE-2 was prevented. Hence, after washing steps the color reaction turned out to be weaker compared to a RBD-antibody-free serum sample. According to the manufacturer, it is assumed from an inhibition of 20% that VNA are present [22].

Second, dilutions of each serum were tested in triplicate in a laboratory-developed Vero cell-based neutralisation assay (cVNT). This 96-well format cVNT uses two own SARS-CoV-2 isolates (alpha, VOC B.1.1.7; delta, VOC B.1.617.2) obtained after cultivation of clinical samples in Vero cells as described previously [26]. A dilution >1:10 was considered likely protective [25, 22].

### Data evaluation and statistical calculations

Data were statistically analysed by help of the GraphPad Prism version 9.1.2 software (GraphPad Software, San Diego, CA, USA). In most cases, the Kruskal-Wallis test, an adjusted, non-paired and non-parametric test was applied. The Wilcoxon test, a paired non-parametric test, was chosen to compare the mean VNA titre differences against VOC B.1.1.7 and VOC B.1.617.2. The level of significance was generally set at 0.05. Furthermore, we calculated the Pearson correlation coefficient r to demonstrate the linear correlation between separate data sets. We used a simple logistic regression to determine the probability of detecting VNA with our cVNT as a function of the anti-SARS-CoV-2 IgG or sVNT results.

## Results

### Composition of the study groups

This study included 59 individuals. Nine and eight of them received a homologous immunisation with AZD1222 or BNT162b2, respectively, while 42 received a heterologous vaccination. The composition of the study groups including median age, age range, gender and median time of blood collection in days is shown in Table 1.

After the first immunisation, all individuals who received an mRNA vaccine developed anti-(trimeric)-S and anti-RBD-IgG. The vaccinees who received the AZD1222 had a prevalence of 55.3 % (anti-S IgG), 76.3% (anti-trimeric S IgG) and 94.7% (anti-RBD IgG), respectively. After administration of the second dose, the IgG prevalence reached 100% in all groups. The mean anti-SARS-CoV-2 IgG titres varied between 39.9 BAU/ml (anti-S IgG; first vaccination in the AZD1222/mRNA group) to 6584 BAU/ml (anti-trimeric S IgG; second vaccination in the BNT162b2/BNT162b2 group). After the second vaccine dose, an increase in mean anti-SARS-CoV-2 IgG titres was observed in all three study groups and in all assays. Compared to the mean anti-SARS-CoV-2 IgG titres after the vector vaccine AZD1222 was administered twice, the corresponding titres were 7 to 10 times higher after a heterologous vaccination and even 10 to 15 times higher according to the homologous BNT162b2 vaccination scheme (Figure 1).

**Figure 1:**
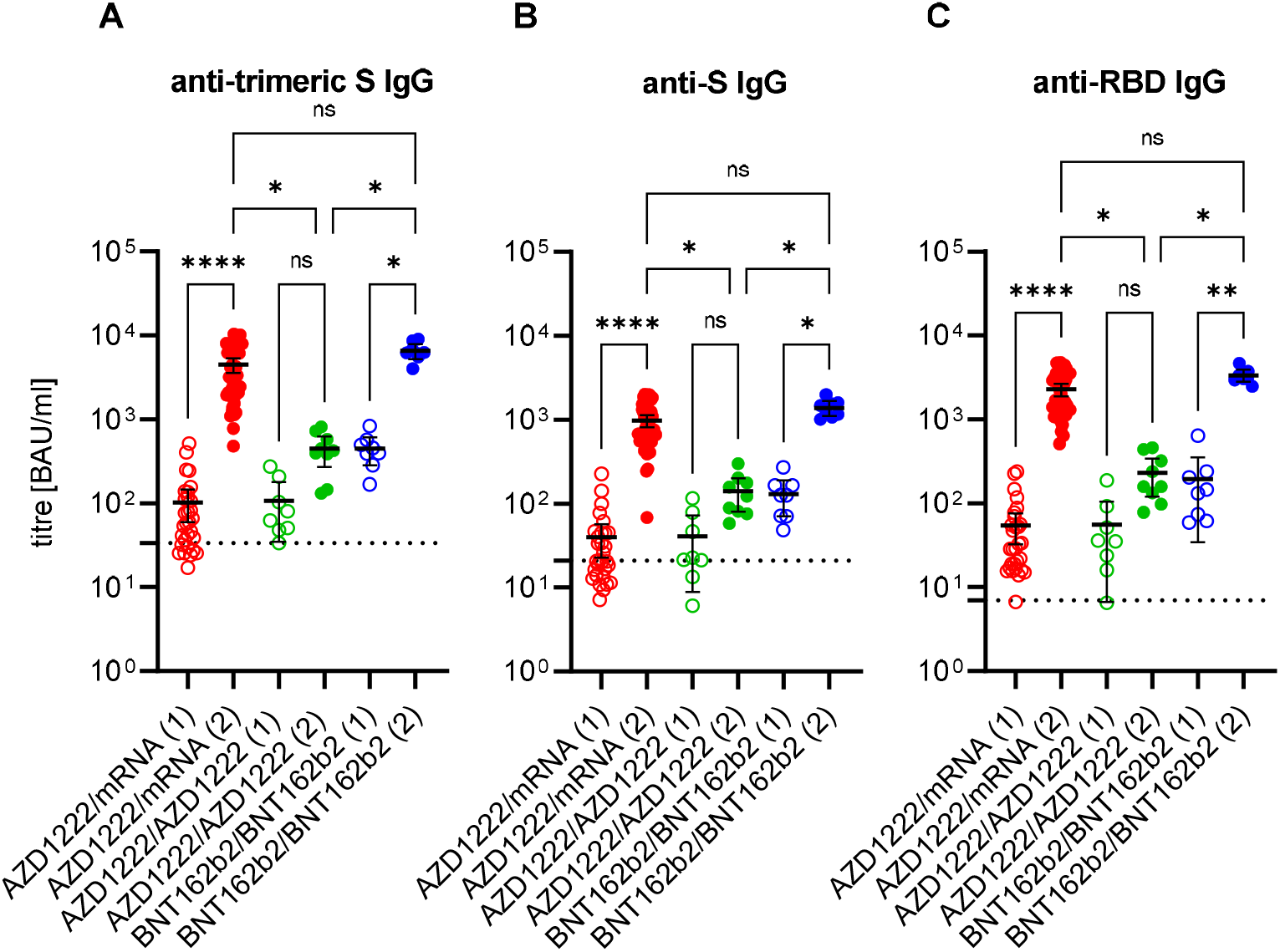
Anti-SARS-CoV-2 immunoglobulin G (IgG) response in Binding Antibody Units (BAU) per milliliter (ml) after first (1; empty circles) and second (2; filled circles) immunisation with the vector vaccine AZD1222 or the messenger ribonucleic acid (mRNA)-based vaccines BNT162b2 or mRNA-1273. The cut-offs for positivity (including borderline results) of anti-trimeric spike (S) IgG assay **(A)**, of the anti-S IgG assay **(B)**, and of the anti-receptor binding domain (RBD) IgG assay **(C)**, respectively, are marked by dashed lines. Ns: non-significant; *: p < 0.05; **: p < 0.01; ****: p < 0.0001 (Kruskal-Wallis test).

Next, we examined the development of anti SARS-CoV-2 IgG avidity in an immunoblot. After the first vaccination, the mean avidity index was in the low to intermediate range. In contrast, high avidities were consistently observed after administration of the second vaccine dose (Figure 2). None of the vaccinees showed an IgG reactivity against the NP in the immunoblot (data not shown).

**Figure 2:**
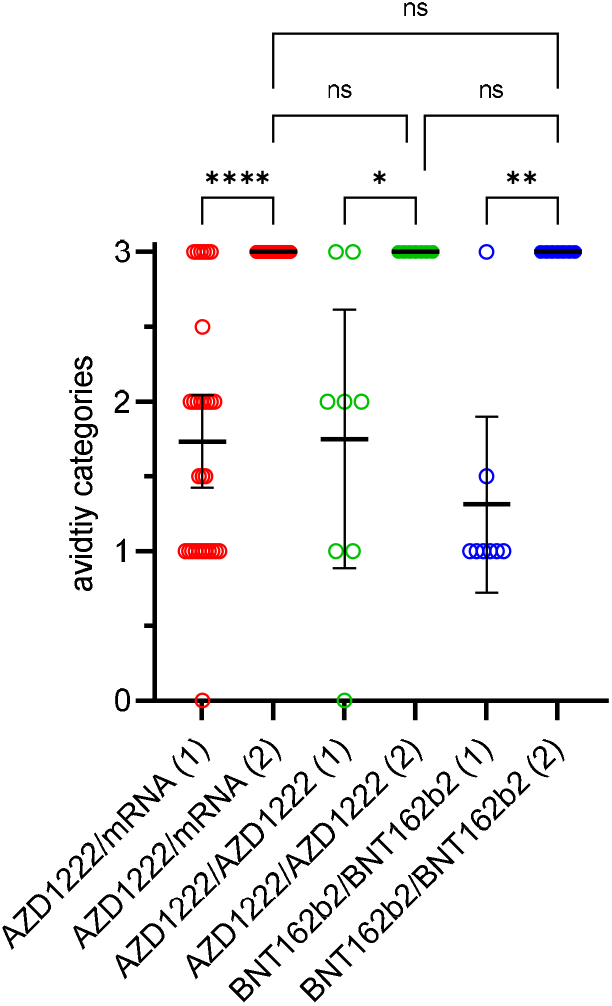
Development of anti-SARS-CoV-2 immunoglobulin G avidities after first (1; empty circles) and second (2; filled circles) immunisation with the vector vaccine AZD1222 or the messenger ribonucleic acid (mRNA)-based vaccines BNT162b2 or mRNA-1273. The measured IgG avidities were assigned to the three categories low (1), intermediate (2) and high (3) and a mean avidity index was calculated per individual serum. Ns: non-significant; *: p < 0.05; **: p < 0.01; ****: p < 0.0001 (Kruskal-Wallis test).

The virus-neutralising properties of the sera were examined with two different assays. A so-called surrogate neutralisation test was used to investigate the extent to which the anti-RBD antibodies that may be present in the serum are able to prevent the binding of this S-protein subunit to the human receptor ACE2. In addition, a laboratory-developed virus-neutralisation test was used, which is based on a VOC B.1.1.7 strain as the antigen. While the majority of the sera in the sVNT were already to be regarded as SARS-CoV-2 neutralising after the first vaccination, these samples did not yet show any corresponding properties in the cVNT. With both methods, however, an increase in the level of VNA could be detected after a second vaccination. There were also marked differences between the three vaccination groups, both in the degree of inhibition (sVNT) and in the level of VNA titres (cVNT). Vaccinees who had received the vector vaccine only had 13- and 11-fold lower geometric mean VNA titres (1:47) compared to individuals immunised heterologously with AZD1222/mRNA (1:608) or homologously with BNT162b2 (1:538), respectively. However, the mean % inhibition of sVNT reached about the same level in all three groups (Figure 3). The quantitative results of the anti-SARS-CoV-2 IgG assays showed an almost perfect correlation to the VNA titres using VOC B.1.1.7 as antigen in the cVNT (Pearson correlation coefficient of 0.89 to 0.91; Figure 4 A-C). In addition, from a certain anti-SARS-CoV-2 IgG titer, it could be assumed with a high degree of probability that VNA are present. The anti-SARS-CoV-2 IgG BAU/ml titres required for this differed between the three assays, but were all in the three-digit range (Figure 4 D-F). The correlation between the measured values of cVNT (VOC B.1.1.7 as antigen) and sVNT was somewhat less pronounced (Pearson correlation coefficient r of 0.84). It is noticeable that especially non-neutralising sera (≤1:10) were overestimated in the sVNT. This is evidenced by the fact that the cut-off set by the manufacturer was only associated with a 4 % probability of detecting VNA in our cVNT (Figure 5).

**Figure 3:**
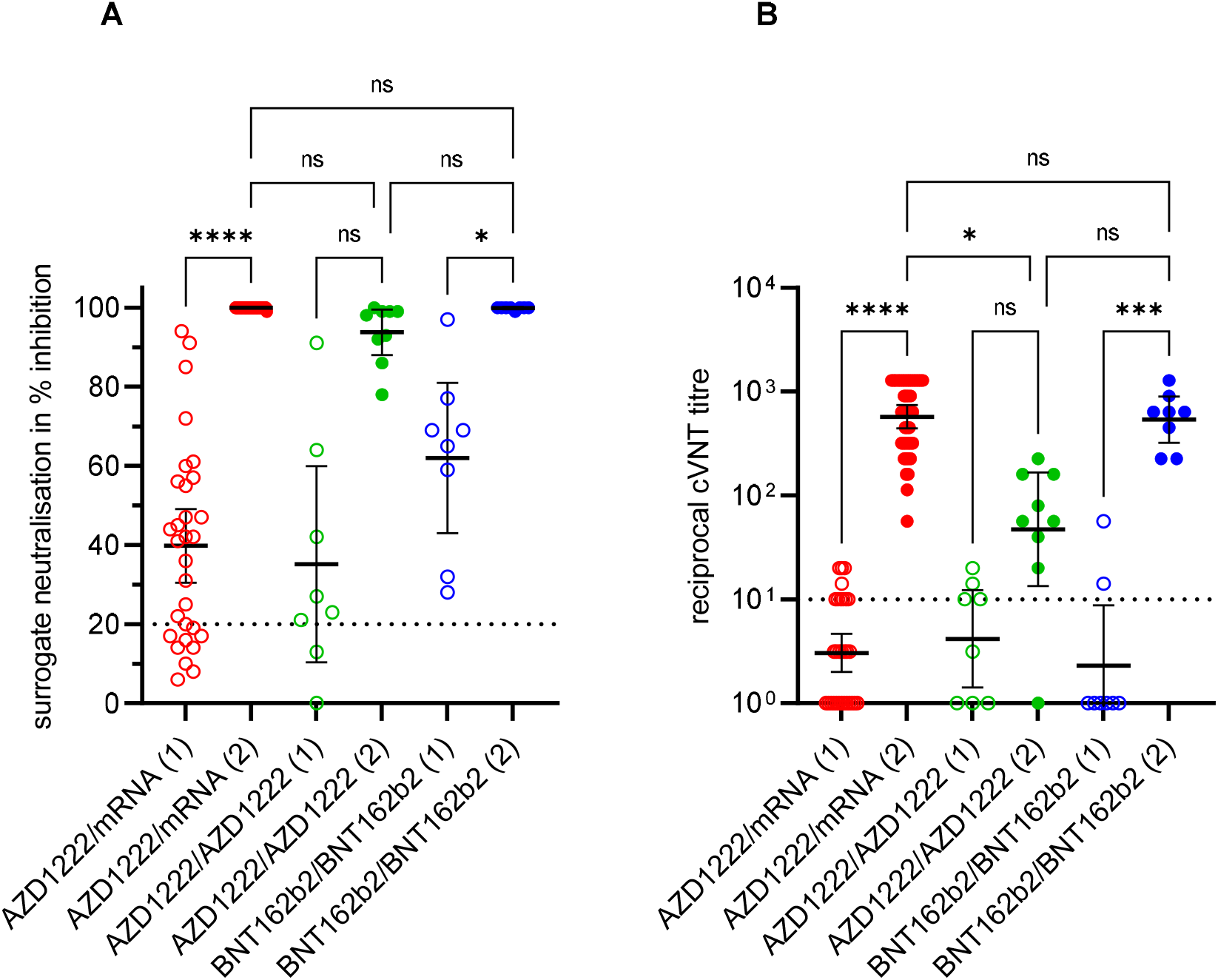
Development of SARS-CoV-2 neutralising antibodies (VNA) after first (1; empty circles) and second (2; filled circles) immunisation with the vector vaccine AZD1222 or the messenger ribonucleic acid (mRNA)-based vaccines BNT162b2 or mRNA-1273. A surrogate neutralisation assay **(A)** and a Vero-cell based virus-neutralisation test (cVNT) using the SARS-CoV-2 variant of concern B.1.1.7 (alpha) strain **(B)** were applied to measure the VNAs. The assay cut-offs are indicated by dashed lines. Ns: non-significant; *: p < 0.05; **: p < 0.01; ****: p < 0.0001 (Kruskal-Wallis test).

**Figure 4:**
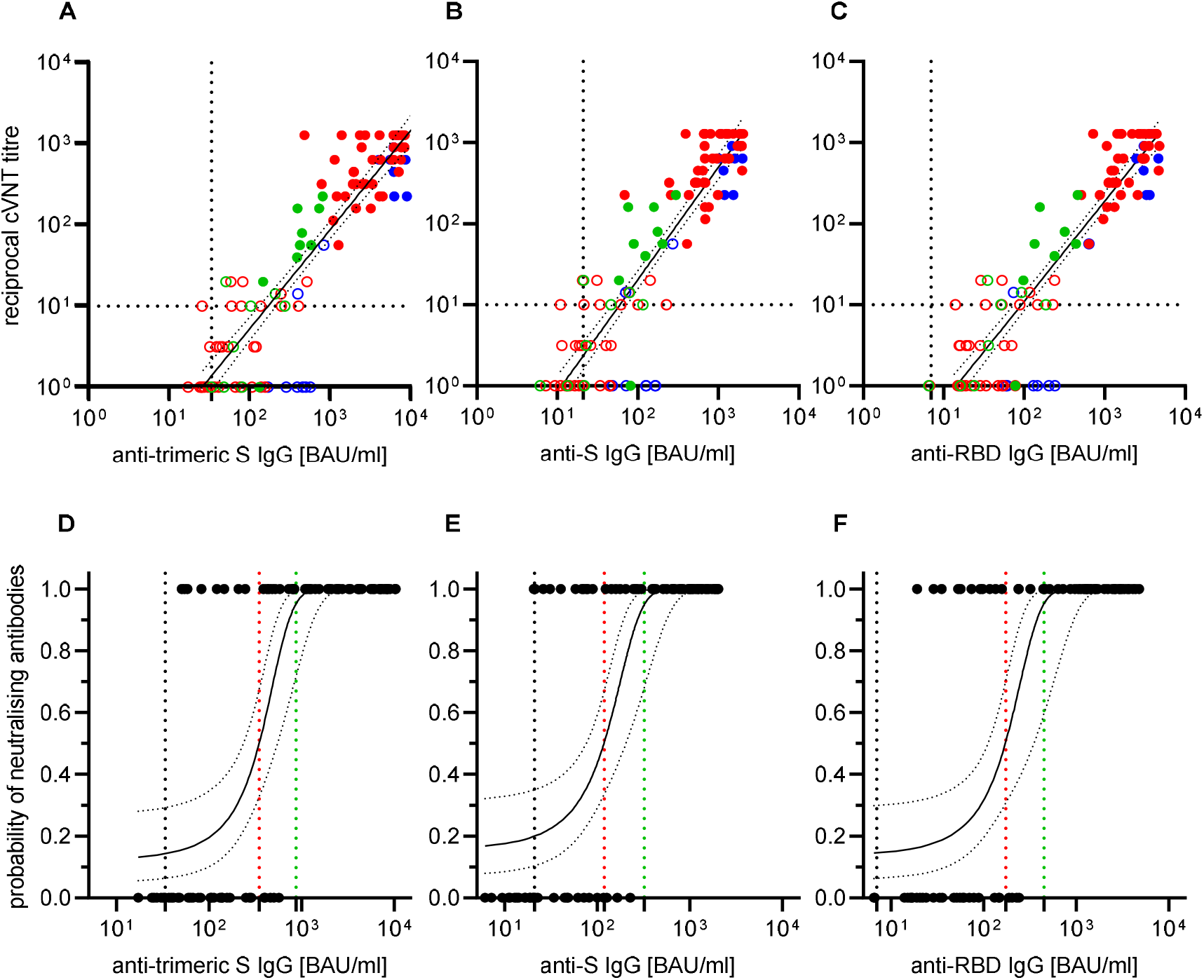
Anti-SARS-CoV-2 immunoglobulin G (IgG) response in Binding Antibody Units (BAU) per milliliter (ml) after first (open circles) and second (filled circles) immunisation with the vector vaccine AZD1222 (green), the messenger ribonucleic acid (mRNA)-based vaccine BNT162b2 (blue) and after a heterologous vaccination scheme, starting with AZD1222, followed by an mRNA-based vaccine boost (BNT162b2 or mRNA-1273; red) with regard to the detection of virus-neutralising antibodies (VNA). The latter were measured in a Vero cell-based neutralisation test (cVNT) using the SARS-CoV-2 variant of concern B.1.1.7 (alpha). Cut-off values for positivity of the anti-trimeric spike (S) IgG assay **(A)**, anti-S IgG assay **(B)** and anti-receptor binding domain (RBD) IgG assay **(C)**, respectively, and the cVNT cut-off value for presence of VNA are indicated by dashed lines. The Pearson correlation coefficients r of log(reciprocal titre) were calculated with 0.89, 0.89, and 0.91, respectively. The probability of detecting VNA at a given BAU/ml in the anti-SARS-CoV-2 IgG assays was calculated by logistic regression **(D-F)**: VNA were present in 95% of samples when titres of 886 BAU/ml (anti-trimeric S IgG), 323 BAU/ml (anti-S IgG), and 448 BAU/ml (anti-RBD IgG) were measured (green dashed lines; 95% confidence intervals 59.4 to 99.6%). Vertical black dashed lines represent the threshold values set by the manufacturers of the antibody assay; red dashed lines represent the BAU/ml values (anti-trimeric S IgG: 350 BAU/ml; anti-S IgG: 119 BAU/ml; anti-RBD IgG: 174 BAU/ml) with a 50% probability of VNA detection.

**Figure 5:**
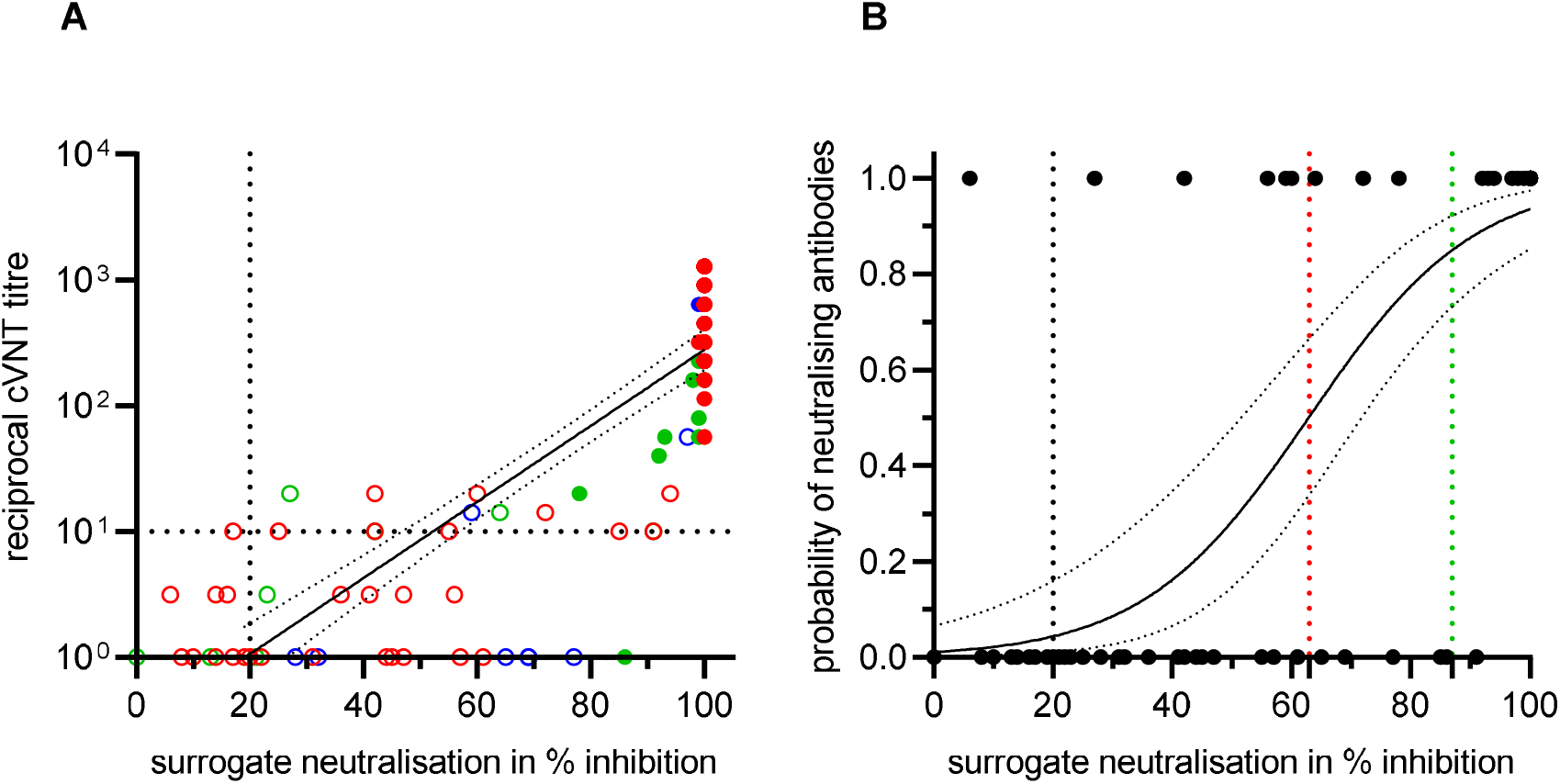
Correlation of the surrogate neutralisation test (sVNT) results with results obtained by the laboratory developed Vero-cell based virus-neutralisation test (cVNT) using a B.1.1.7 strain as antigen **(A)**. The Pearson correlation coefficient r of log(reciprocal titre) was calculated with 0.84; empty circles; first vaccination; filled circles: second vaccination; red: heterologous vaccination with AZD1222/mRNA; green: homologous vaccination with AZD1222; blue: homologous vaccination with BNT162b2. Probability of detecting virus-neutralising antibodies (VNA) with the cVNT at a given % inhibition of sVNT calculated by logistic regression **(B)**; e.g. at 20% inhibition (black dashed line), 63 % inhibition (red dashed line), and at 87% inhibition of sVNT (green dashed line), the probabilities of detecting VNA with cVNT are 4% (95% confidence interval (CI) 1 %-16 %), 50 % (95 % CI 34 %-66 %) and 85% (95 % CI 73 %-92 %), respectively.

A subset of age-and gender matched sera (Table 1) obtained after the second immunisation were also tested in the cVNT for presence of VNA against VOC B.1.617.2 (delta). In comparison to B.1.1.7, all three vaccination groups exhibited significantly lower mean VNA titres (AZD1222/BNT162b2: 838.0 vs. 89.8, 9.3-fold lower; AZD1222/AZD1222: 47.3 vs. 9.0, 5.3-fold lower; BNT162b2/BNT162b2: 538.2 vs. 160.0, 3.4-fold lower) when B.1.617.2 is used as the antigen. If the vaccination schedule is used as a basis, those vaccinated homologously with the vector vaccine had significantly lower mean VNAs than those homologously vaccinated with BNT162b2. This difference was also found for the heterologous vaccination regime. In contrast, the mean VNAs between those vaccinated homologously with BNT162b and those vaccinated heterologously (AZD1222/BNT162b2) exhibited no measurable difference (Figure 6).

**Figure 6:**
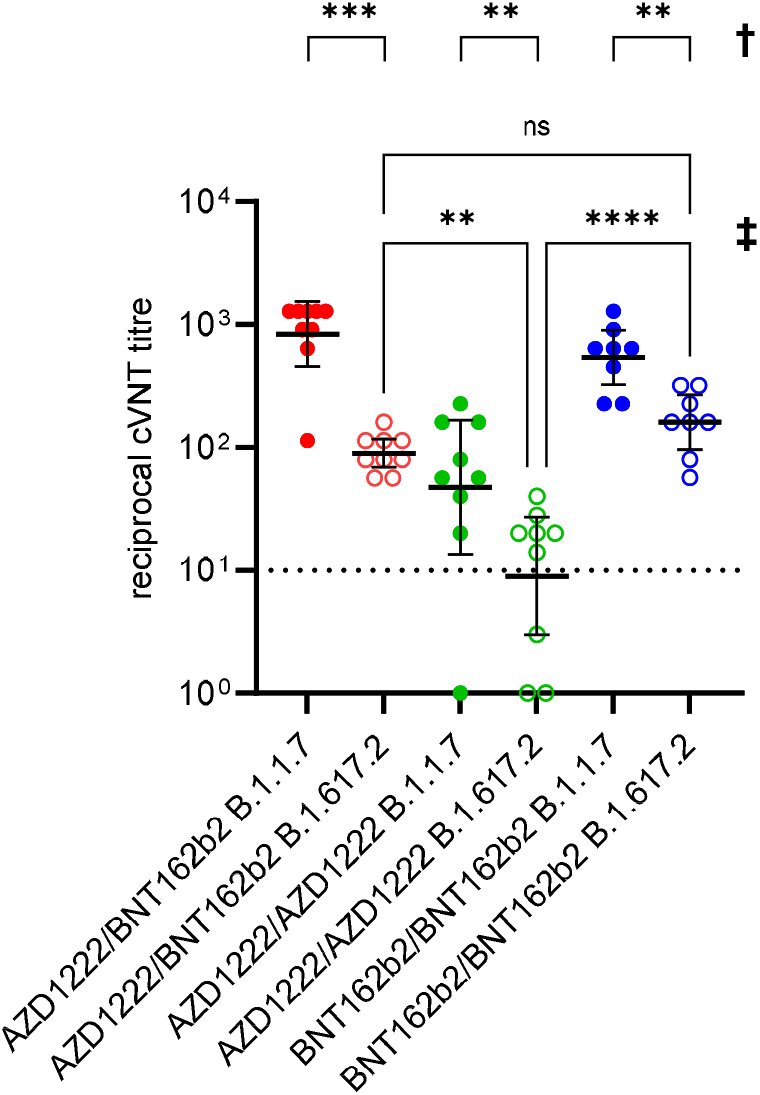
Presence of virus-neutralising antibodies (VNA) against the SARS-CoV-2 variants of concern B.1.1.7 (alpha) and B.1.617.2 (delta) after the second immunisation. Sera from 26 age and gender matched individuals who received a heterologous (AZD1222/BNT162b2, n=9) or a homologous vaccination scheme (AZD1222/AZD1222, n=9; BNT162b2/BNT162b2, n=8) were tested (see Table 1). An individual VNA titre > 1:10 was defined as neutralising in our Vero cell-based virus neutralisation test (cVNT). **†** The significance of the mean VNA titre differences against B.1.1.7 and B.1.617.2 were calculated using the Wilcoxon test (**: p < 0.01; ***: p < 0.001). **‡** Comparison of the mean VNA titre differences achieved with different immunisation schemes against B.1.617.2 (Kruskal-Wallis test; ns: not significant; *: p < 0.05; ****: p < 0.0001).

## Discussion

Due to the rare but serious side effects after administration of the vector vaccine, in spring 2021 the STIKO recommended that people under the age of 60 should complete vaccinations that had already been started with a vector vaccine with an mRNA vaccine [9, 10]. This recommendation was extended on July 1st, 2021 to all who had already received a primary vaccination with AZD1222 [12]. In the first quarter of this year, however, only a few animal experimental data were available on the immunological outcome of the proposed heterologous vaccination scheme [13, 14]. Several studies have now appeared on the immunogenicity of the heterologous vaccination scheme in humans [16, 21, 17-20].

In this report, we compared the development of the humoral immune response after homologous and heterologous vaccination with different methods. To the best of our knowledge, we are one of the first research groups to investigate the anti-delta VOC neutralising effect of sera after completing the heterologous immunisation regime.

After the first vaccination, the majority of individuals developed anti-trimeric-S, -S, and -RBD IgG antibodies, respectively. However, their titres varied between the three study groups. The results are in line with our previous study [22]. It is evident that the anti-SARS-CoV-2 IgG titres are not comparable between the three assays either. This is probably due to the different antigen preparations. In this respect, from our point of view, titre comparisons are only possible if they have been measured with the same assay.

The second immunisation resulted in higher titres in all three groups. It was noticeable that significantly higher anti-SARS-CoV-2 IgG titres were detected after a second vaccination with an mRNA vaccine than after the vector vaccine AZD1222 was administered again. The increase in anti-S and anti-RBD IgG titres after a second vaccination with an mRNA vaccine confirms our previous study. Due to the recommended vaccination interval of 10 to 12 weeks, we did not yet have any data on the development of the SARS-CoV-2-specific IgG antibodies after the second administration of a vector vaccine [22]. The lack of NP-specific IgG antibodies in all vaccinees can be interpreted as an indication that they had no SARS-CoV-2 infection and were therefore to be regarded as immunologically naive before immunisation [22]. It is known that vaccinations lead to particularly high anti-SARS-CoV-2 IgG titres in convalescents [27]. For these individuals, the recommendation is that they should receive a vaccine dose about six months after they have been infected [10].

After the first vaccination nearly all vaccinees exhibited only low to intermediate avid anti-SARS-CoV-2 IgG, while after the second vaccine dose IgG of high avidity appeared in all cases. These results confirm and expand the existing knowledge on the development of highly avid anti-SARS-CoV-2 IgG after a second vaccination with an mRNA vaccine [28]. In line with this, VNA > 1:10 against the previously prevalent SARS-CoV-2 VOC B.1.1.7 (alpha) were observed after second immunisation which confirms our recent study [22]. Marked differences in mean VNA titres were observed between individuals re-vaccinated with an mRNA vaccine and those re-vaccinated with a vector vaccine which is in accordance to animal experiments [13]. The anti-SARS-CoV-2 IgG titres obtained with three preparations of the viral S-protein correlated well with the presence and level of VNA using B.1.1.7 as the antigen in our cVNT. This observation is in line with a recently published study from Finland [29] and suggests that these standardised and easy-to-perform commercial tests are useful for demonstrating vaccination success. Anti-SARS-CoV-2 IgG titres that indicate the presence of VNA cannot, however, be defined across the board. These depend on the cVNT and the viral antigen used in it. Basically, the sVNT also showed the titre increase and evaluated all sera as virus-neutralising after the second vaccination. However, it is again noticeable that sera that are not or only weakly SARS-CoV-2-neutralising in the in-house cVNT are already evaluated as neutralising in the sVNT. This result supports our previous suggestion to raise the cut-off of the sVNT [22]. Compared to the admittedly very conservative in-house cVNT, a cut-off of over 80% would be desirable. Due to the lack of standardisation of the widespread cVNTs for the detection of VNA against SARS-CoV-2, this recommendation only applies to our laboratory and cannot be generalised.

From our point of view, a particularly interesting point is the significantly reduced capacity for neutralising the SARS-CoV-2 delta variant (VOC B.1.617.2) *in vitro* using a cVNT. We observed this in all 26 age and gender matched individuals regardless of the immunisation regime. In addition, three of nine vaccinees who had received two doses of AZD1222 presented low or undetectable VNA against this VOC, which is considered to be 60% more transmissible than alpha [30]. The SARS-CoV-2 delta VOC is known to have accumulated a number of mutations in the S-protein. These enable continued good binding to the cellular ACE2 receptor, but at the same time lead to the viral S-protein being less efficiently recognised by antibodies [30]. The significantly lower VNA titres compared to the alpha VOC, which we and others [30, 31] observed, corroborate the suspicion of an immunescape of the delta VOC. Based on our VNA data for B.1.1.7, it can be assumed that a single vaccination is not sufficient to induce measurable VNA against B.1.617.2.

The results presented by us on the antibody response after heterologous SARS-CoV-2 vaccination are consistent with the few available clinical studies [17, 19, 21, 16, 18, 20]. In June 2021, a randomised study from Spain has already demonstrated that the heterologous vaccination scheme is suitable for generating a robust immune response. Unfortunately, this very extensive work did not include a control group of individuals who received two immunisations with the vector vaccine [21]. A recent preprint reports significant higher anti-S antibody titres in a group of 26 individuals who first received an AZD1222 vaccination followed by re-vaccination with BNT162b2 compared to 14 individuals that were vaccinated twice with BNT162b2. However, these results were obtained with a total antibody assay which does not discriminate between IgG and acute phase immunoglobulin M. Furthermore, markedly higher VNA titres were observed in the AZD1222/BNT162b2 group using chimeric vesicular stomatitis viruses carrying the S-proteins of SARS-CoV-2 VOCs B.1.1.7 (alpha) or B.1.351 (beta), respectively, as antigens. However, data after homologous vaccination with the vector vaccine are missing in this preliminary report [16]. In another investigation [18], differences in the anti-S IgG and VNA titres were not observed after heterologous vaccination (AZD1222/mRNA) or after homologous vaccination with an mRNA vaccine. In contrast, both parameters were significantly lower after homologous vaccination with the vector vaccine. These results are in agreement with our data even if only one IgG assay and one sVNT were used by this research group [18]. The results of a single-blind randomised British study, in which the four possible vaccine combinations of AZD1222 and BNT162b2 were compared with one another, are very interesting and promising. These researchers report about 9-fold higher mean anti-S IgG titres in sera from heterologous AZD1222/BNT162b2 vaccinees compared to individuals immunised twice with AZD1222 [19] which is largely in accordance to our results. In a study from Sweden, markedly higher anti-S and anti-RBD IgG titres were observed after heterologous vaccination (AZD1222/mRNA-1273) compared to the homologues AZD1222 scheme. In line with this the occurrence of significantly higher VNA titres both against a wild-type SARS-CoV-2 and against a VOC B.1.351 (beta) strain were demonstrated for these vaccinees [20]. One investigation reports the development of high IgG avidity after completion of a homologous (AZD1222/AZD1222; BNT162b2/BNT162b2) or heterologous SARS-CoV-2 (AZD1222/BNT162b2) immunisation scheme. While there were no qualitative differences in the development of IgG avidity between both groups, the AZD1222/BNT162b2 vaccinees developed a significantly higher relative avidity index [17]. In our assay we cannot investigate such gradual differences. Furthermore, the authors report markedly lower anti-SARS-CoV-2 IgG and VNA (against VOCs alpha and beta) titres in the AZD1222/AZD1222 group [17].

It is not yet sufficiently clear why homologous vaccination with the AZD1222 vector vaccine leads to lower anti-SARS-CoV-2 IgG and VNA titres. A possible explanation could be the immune response to the adenovirus vector backbone (so called “antivector immunity” [3]).

The work presented by us contributes to a better understanding of the humoral immunogenicity of the heterologous SARS-CoV-2 vaccination regimen. With various assays we monitored the development of anti S-specific IgG antibodies and make statements about their binding-strength as an expression of maturity. In addition, with a commercial and an in-house test, we showed that VNA can be detected after the second vaccination and that VNA titres vary in dependence of the viral antigen.

Important limitations of our report are (i) the heterogeneity of the study groups, (ii) the small group size of individuals who received a homologous vaccination scheme, (iii) the subjects, who are predominantly in younger to middle adulthood, (iv) the lack of information on the durability of the detected antibodies, and (v) the missing consideration of cellular and innate immunity after immunisation. Therefore, among other things, no statements can be made from our data about the need for further booster vaccinations. In addition, no better protection against SARS-CoV-2 infections can be derived from the level of the antibody titre *per se*.

In summary, the administration of a vector vaccine followed by an mRNA vaccine boost resulted in a strong humoral immune response, comparable to that after two immunisations with an mRNA vaccine. Regardless of the vaccination scheme, all individuals developed highly avid anti-SARS-CoV-2 IgGs. Even if almost all vaccinees presented VNA after the second immunisation, the generally lower titres indicate a partial immune escape of the delta VOC.

## Data Availability

The data presented in this study are available on request from the corresponding author.

## Availability of data

The data presented in this study are available on request from the corresponding author.

## Ethics approval and consent to participate

The study was conducted according to the guidelines of the Declaration of Helsinki, and approved by the Ethics Committee of the medical faculty of the Christian-Albrechts-Universität zu Kiel, Kiel, Germany (D467/20, 16.04.2020; amendment 02.02.2021). Informed consent was obtained from all subjects involved in the study.

## Consent for publication

Not applicable.

## Author Contributions

AK, FN, and RR had full access to all the data in the study and take responsibility for the integrity of the data and the accuracy of the data analysis. Concept and design: AK. Acquisition, analysis, or interpretation of the data: all authors. Drafting of the manuscript: AK, HF and RR. Critical revision of the manuscript for important intellectual content: all authors. All authors have read and agreed to the published version of the manuscript.

## Acknowledgments

The authors would like to thank all volunteers for participating in the study.

## References

1. Hu B, Guo H, Zhou P, Shi ZL. Characteristics of SARS-CoV-2 and COVID-19. Nat Rev Microbiol. 2021;19(3):141–54. doi:10.1038/s41579-020-00459-7.

2. Klasse PJ, Nixon DF, Moore JP. Immunogenicity of clinically relevant SARS-CoV-2 vaccines in nonhuman primates and humans. Sci Adv. 2021;7(12). doi:10.1126/sciadv.abe8065.

3. Sadarangani M, Marchant A, Kollmann TR. Immunological mechanisms of vaccine-induced protection against COVID-19 in humans. Nat Rev Immunol. 2021;21(8):475–84. doi:10.1038/s41577-021-00578-z.

4. Krammer F. SARS-CoV-2 vaccines in development. Nature. 2020;586(7830):516–27. doi:10.1038/s41586-020-2798-3.

5. Cines DB, Bussel JB. SARS-CoV-2 Vaccine-Induced Immune Thrombotic Thrombocytopenia. N Engl J Med. 2021;384(23):2254–6. doi:10.1056/NEJMe2106315.

6. Kyriakidis NC, Lopez-Cortes A, Gonzalez EV, Grimaldos AB, Prado EO. SARS-CoV-2 vaccines strategies: a comprehensive review of phase 3 candidates. NPJ Vaccines. 2021;6(1):28. doi:10.1038/s41541-021-00292-w.

7. Creech CB, Walker SC, Samuels RJ. SARS-CoV-2 Vaccines. JAMA. 2021;325(13):1318–20. doi:10.1001/jama.2021.3199.

8. EU. Safe COVID-19 vaccines for Europeans. 2021. https://ec.europa.eu/info/live-work-travel-eu/coronavirus-response/safe-covid-19-vaccines-europeans_en. Accessed 12 May 2021.

9. STIKO. Stellungnahme der Ständigen Impfkommission zum Zeitpunkt der Gabe eines mRNA-Impfstoffs nach Erstimpfung mit AstraZeneca Vaccine (Vaxzevria) bei <60-Jährigen. 2021. https://www.rki.de/DE/Content/Kommissionen/STIKO/Empfehlungen/Stellungnahme-Impfabstand.html. Accessed 12 May 2021.

10. RKI. Mitteilung der Ständigen Impfkommission beim Robert Koch-Institut Beschluss der STIKO zur 7. Aktualisierung der COVID-19-Impfempfehlung und die dazugehörige wissenschaftliche Begründung. Epidemiologisches Bulletin. 2021;25:3–13.

11. Rambaut A, Holmes EC, O’Toole A, Hill V, McCrone JT, Ruis C et al. A dynamic nomenclature proposal for SARS-CoV-2 lineages to assist genomic epidemiology. Nat Microbiol. 2020;5(11):1403–7. doi:10.1038/s41564-020-0770-5.

12. STIKO. Mitteilung der STIKO zur COVID-19-Impfung: Impfabstand und heterologes Impfschema nach Erstimpfung mit Vaxzevria (1.7.2021). 2021. https://www.rki.de/DE/Content/Kommissionen/STIKO/Empfehlungen/PM_2021-07-01.html. Accessed 2 July 2021.

13. Spencer AJ, McKay PF, Belij-Rammerstorfer S, Ulaszewska M, Bissett CD, Hu K et al. Heterologous vaccination regimens with self-amplifying RNA and adenoviral COVID vaccines induce robust immune responses in mice. Nat Commun. 2021;12(1):2893. doi:10.1038/s41467-021-23173-1.

14. He Q, Mao Q, An C, Zhang J, Gao F, Bian L et al. Heterologous prime-boost: breaking the protective immune response bottleneck of COVID-19 vaccine candidates. Emerg Microbes Infect. 2021;10(1):629–37. doi:10.1080/22221751.2021.1902245.

15. Shaw RH, Stuart A, Greenland M, Liu X, Nguyen Van-Tam JS, Snape MD et al. Heterologous prime-boost COVID-19 vaccination: initial reactogenicity data. Lancet. 2021;397(10289):2043–6. doi:10.1016/S0140-6736(21)01115-6.

16. Groß R, Zanoni M, Seidel A, Conzelmann C, Gilg A, Krnavek D et al. Heterologous ChAdOx1 nCoV-19 and BNT162b2 prime-boost vaccination elicits potent neutralizing antibody responses and T cell reactivity. medRxiv. 2021:2021.05.30.21257971. doi:10.1101/2021.05.30.21257971.

17. Hillus D, Schwarz T, Tober-Lau P, Vanshylla K, Hastor H, Thibeault C et al. Safety, reactogenicity, and immunogenicity of homologous and heterologous prime-boost immunisation with ChAdOx1 nCoV-19 and BNT162b2: a prospective cohort study. Lancet Respir Med. 2021. doi:10.1016/S2213-2600(21)00357-X.

18. Schmidt T, Klemis V, Schub D, Mihm J, Hielscher F, Marx S et al. Immunogenicity and reactogenicity of heterologous ChAdOx1 nCoV-19/mRNA vaccination. Nature Medicine. 2021. doi:10.1038/s41591-021-01464-w.

19. Liu X, Shaw RH, Stuart ASV, Greenland M, Aley PK, Andrews NJ et al. Safety and immunogenicity of heterologous versus homologous prime-boost schedules with an adenoviral vectored and mRNA COVID-19 vaccine (Com-COV): a single-blind, randomised, non-inferiority trial. Lancet. 2021. doi:10.1016/S0140-6736(21)01694-9.

20. Normark J, Vikstrom L, Gwon YD, Persson IL, Edin A, Bjorsell T et al. Heterologous ChAdOx1 nCoV-19 and mRNA-1273 Vaccination. N Engl J Med. 2021. doi:10.1056/NEJMc2110716.

21. Borobia AM, Carcas AJ, Perez-Olmeda M, Castano L, Bertran MJ, Garcia-Perez J et al. Immunogenicity and reactogenicity of BNT162b2 booster in ChAdOx1-S-primed participants (CombiVacS): a multicentre, open-label, randomised, controlled, phase 2 trial. Lancet. 2021;398(10295):121–30. doi:10.1016/S0140-6736(21)01420-3.

22. Neumann F, Rose R, Rompke J, Grobe O, Lorentz T, Fickenscher H et al. Development of SARS-CoV-2 Specific IgG and Virus-Neutralizing Antibodies after Infection with Variants of Concern or Vaccination. Vaccines (Basel). 2021;9(7):700. doi:10.3390/vaccines9070700.

23. Bonelli F, Blocki FA, Bunnell T, Chu E, De La OA, Grenache DG et al. Evaluation of the automated LIAISON((R)) SARS-CoV-2 TrimericS IgG assay for the detection of circulating antibodies. Clin Chem Lab Med. 2021;59(8):1463–7. doi:10.1515/cclm-2021-0023.

24. Kristiansen PA, Page M, Bernasconi V, Mattiuzzo G, Dull P, Makar K et al. WHO International Standard for anti-SARS-CoV-2 immunoglobulin. Lancet. 2021;397(10282):1347–8. doi:10.1016/S0140-6736(21)00527-4.

25. Stromer A, Rose R, Grobe O, Neumann F, Fickenscher H, Lorentz T et al. Kinetics of Nucleo-and Spike Protein-Specific Immunoglobulin G and of Virus-Neutralizing Antibodies after SARS-CoV-2 Infection. Microorganisms. 2020;8(10). doi:10.3390/microorganisms8101572.

26. Stromer A, Rose R, Schafer M, Schon F, Vollersen A, Lorentz T et al. Performance of a Point-of-Care Test for the Rapid Detection of SARS-CoV-2 Antigen. Microorganisms. 2020;9(1). doi:10.3390/microorganisms9010058.

27. Frieman M, Harris AD, Herati RS, Krammer F, Mantovani A, Rescigno M et al. SARS-CoV-2 vaccines for all but a single dose for COVID-19 survivors. EBioMedicine. 2021;68. doi:10.1016/j.ebiom.2021.103401.

28. Struck F, Schreiner P, Staschik E, Wochinz-Richter K, Schulz S, Soutschek E et al. Vaccination versus infection with SARS-CoV-2: Establishment of a high avidity IgG response versus incomplete avidity maturation. J Med Virol. 2021. doi:10.1002/jmv.27270.

29. Jalkanen P, Kolehmainen P, Häkkinen HK, Huttunen M, Tähtinen PA, Lundberg R et al. COVID-19 mRNA vaccine induced antibody responses against three SARS-CoV-2 variants. Nature Communications. 2021;12(1):3991. doi:10.1038/s41467-021-24285-4.

30. Planas D, Veyer D, Baidaliuk A, Staropoli I, Guivel-Benhassine F, Rajah MM et al. Reduced sensitivity of SARS-CoV-2 variant Delta to antibody neutralization. Nature. 2021;596(7871):276–80. doi:10.1038/s41586-021-03777-9.

31. Wall EC, Wu M, Harvey R, Kelly G, Warchal S, Sawyer C et al. Neutralising antibody activity against SARS-CoV-2 VOCs B.1.617.2 and B.1.351 by BNT162b2 vaccination. Lancet. 2021;397(10292):2331–3. doi:10.1016/S0140-6736(21)01290-3.

